# IL-8 as a potential link between aging and impaired influenza antibody responses in older adults

**DOI:** 10.1101/2024.11.07.24316936

**Authors:** Huy Quang Quach, Krista M. Goergen, Diane E. Grill, Inna G. Ovsyannikova, Gregory A. Poland, Richard B. Kennedy

## Abstract

**Background:** Antibody responses to MF59-adjuvanted (MF59Flu) and high-dose (HDFlu) influenza vaccines have been well-characterized in older adults, yet corresponding cellular response data remain limited.

**Methods:** Blood samples were collected from 106 MF59Flu recipients and 112 HDFlu recipients before vaccination (Day 0), and on Days 1, 8, and 28 post-vaccination. Antibody responses were assessed on Days 0, 8, and 28 using a hemagglutination inhibition (HAI) assay. Eight pro-inflammatory cytokines and chemokines, including IFN-α2a, IFN-γ, IP-10, MCP-1, MIP-1α, IL-1β, IL-6, IL-8, were quantified from PBMCs collected on Days 0 and 1 following stimulation with live influenza A/H3N2 virus using a multiplex assay. Associations between cytokine/chemokine levels and HAI titers were examined, along with the effect of sex, age, body mass index (BMI), and cytomegalovirus infection status.

**Results:** Vaccine type (MF59Flu or HDFlu), sex, BMI and cytomegalovirus infection did not significantly impact cytokine and chemokine levels. However, age was positively correlated with IL-8 level on Day 1 (r = 0.24, *p* = 0.0003) as well as the change in IL-8 levels from Day 1 to Day 0 (r = 0.16, *p* = 0.021). Notably, the change in IL-8 levels was negatively associated with peak antibody responses at Day 28 (r = −0.15, *p* = 0.026).

**Conclusion:** Our findings underscore IL-8 as a potential link between aging and impaired antibody responses to influenza vaccination in older adults, suggesting that IL-8 inhibition could be a promising molecular intervention to improve immunogenicity and efficacy of influenza vaccines in this high-risk population.

## 1. Introduction

Older adults (≥65 years) are particularly vulnerable to severe influenza illness, accounting for up to 85% of deaths and 70% of hospitalizations related to seasonal flu [1]. This heightened risk is primarily attributed to immunosenescence, the functional decline of T and B cells with age [2]. To address this, MF59-adjuvanted (MF59Flu) and high-dose (HDFlu), two enhanced influenza vaccines, have been recommended for this population. While most studies suggest that the two vaccines offer similar effectiveness in older adults [3–5], some studies favor MF59Flu [6], and others support HDFlu [7]. These discrepancies highlight the challenges of assessing vaccine efficacy and emphasize the need for further research to identify factors that may influence these differential outcomes.

Conventionally, influenza vaccine effectiveness is assessed by measuring the titer of neutralizing hemagglutination inhibiting antibodies or by differentiating influenza-specific memory T cells, with antibody considered as the primary correlate of protection for influenza [8]. However, effective activation of T and B cells is primarily orchestrated by the local cytokine and chemokine environment, which is shaped by the activation of innate immune cells. This cytokine and chemokine milieu can have paradoxical effects on host defense against influenza [9, 10]. On the one hand, cytokines and chemokines can exert antiviral activities by inhibiting influenza virus replication [11, 12] and promoting T and B cell immunity [13, 14]. Conversely, excessive secretion of pro-inflammatory cytokines and chemokines have been linked to severe disease outcomes [15–21]. In response to influenza vaccination, studies have shown that increases in TNF-α response [22] and decreases in IL-8 production [23] are correlated with improved antibody responses in vaccine responders. However, vaccine non-responders and responders had comparable levels of IFN-γ, IL-6, and IL-10 [24]. These mixed results underscore the need for further research to clarify the role of these cytokines and chemokines in influenza immunity. This is especially important in older adults, who are often affected by “inflammaging”, a condition characterized by elevated level of inflammatory markers with age [25, 26].

In this study, we isolated PBMCs from MF59Flu and HDFlu recipients and characterized cytokine and chemokine responses in these cells upon *in vitro* stimulation with influenza virus. Additionally, we examined the potential impact of vaccine type, sex, age, body mass index (BMI), and cytomegalovirus (CMV) infection status on cytokine and chemokine profiles.

## 2. Methods

The methods used in this study are identical or similar to the methods used in our previous publications [27–29].

### 2.1. Study participants

This study was reviewed and approved by the Mayo Clinic Institutional Review Board (IRB# 17-010601) and conducted in accordance with the Declaration of Helsinki. All study participants provided written informed consent prior to enrollment in this study.

A total of 250 participants (≥65 years) were recruited from Olmsted County, MN, and the surrounding areas for this study. Participants were randomly assigned to receive either the MF59-adjuvanted inactivated influenza vaccine (MF59Flu, marketed as Fluad®) or high-dose inactivated influenza vaccine (HDFlu, marketed as Fluzone®). Both vaccines are trivalent, containing three viral strains: i) H1N1 (A/Michigan/45/2015/pdm09-like strain), ii) H3N2 (A/Singapore/INFIMH-16-0019/2016-like strain), and iii) influenza B lineage (Colorado/06/2017-like Victoria lineage strain). Vaccination histories and health status were documented for all participants. Additionally, body mass index (BMI) was calculated from height and weight data collected on all study participants. Participants who exhibited symptoms associated with influenza infection during the study period were excluded from data analysis.

### 2.2. Processing blood samples

Blood samples were collected from each participant at four timepoints: prior to the receipt of vaccination (Day 0) and on Days 1, 8, and 28 after vaccination. Serum and peripheral blood mononuclear cells (PBMCs) were isolated from blood samples using a standard protocol established in our laboratory [28]. Both serum samples and PBMCs were stored at −80 °C until further analysis.

### 2.4. Hemagglutination inhibition assay

We performed a hemagglutination inhibition (HAI) assay against the influenza A/H3N2 virus (A/Singapore/INFIMH-16-0019/2016/H3N2 strain) using sera collected at Days 0, 8, and 28. HAI results from Day 0 and Day 28 sera from this study cohort were partially summarized in our earlier publication [27]. In the current study, we investigated the associations between cytokine and chemokine concentrations secreted by PBMCs sampled at Day 0 and Day 1 and the HAI titers at Days 0, 8, and 28. The coefficient of variation for the HAI assay in our laboratory was 2.9% [30].

### 2.5. Cytokine and chemokine assays

Cytokine and chemokine secretion was quantified using multiplex electrochemiluminescence-based kits (Meso Scale Diagnostics, Rockville, MD). Briefly, 2×10^5^ live PBMCs sampled at Day 0 and Day 1 were plated in triplicates in each well of a 96-well round bottom plate. The cells were then incubated under one of following conditions: i) RPMI media alone (unstimulated), ii) live influenza A/H3N2 virus (A/Singapore/INFIMH-16-0019/2016-like strain; multiply of infection or MOI=0.5) in RPMI media, or iii) phytohemagglutinin (PHA; 10 μg/mL) in RPMI media. After an 18-hour incubation at 37 °C with 5% CO_2_, the cell culture supernatants were collected and stored at −80°C until for batched measurements. For the multiplex assay, supernatants were diluted 2-fold with assay diluent and assayed for the following cytokines and chemokines: IFN-α2a, IFN-γ, IL-1β, IL-6, IL-8, interferon gamma-induced protein (IP)-10, monocyte chemoattractant protein (MCP)-1, and macrophage inflammatory protein (MIP)-1α. The assay was conducted according to the manufacturer’s protocol. The plates were read, and the results were analyzed at the Mayo Clinic Immunochemical Core Laboratory. According to the manufacturer, the coefficient of variation for each analyte was <20%.

### 2.6. Measurement of cytomegalovirus (CMV) IgG

Serum level of CMV IgG was measured using a Bio-Rad CMV IgG EIA kit (catalog no. 25177), with results reported as a sample index. The sample index was then used to categorize results as either CMV seronegative or seropositive, following the manufacturer’s instruction. The assay had an intra-assay coefficient of variation (CV) of 8.3% and inter-assay CV of 9.5%, according to the manufacturer.

### 2.7. Statistical analysis

The Wilcoxon rank-sum test was used to assess the differences in age, weight, height, BMI, and analyte concentrations between two vaccine groups (MF59Flu and HDFlu). The differences in categorical variables (sex, ethnicity, and race) were evaluated by Pearson’s Chi-squared test. The association between analyte concentrations and variables such as age, BMI, and HAI titer was assessed by Spearman’s rank correlation.

## 3. Results

### 3.1. Characteristics of study cohort

The demographic characteristics of this cohort were partially reported in Table 1 of our earlier publication [27, 29]. Briefly, we recruited 250 older adults (≥65 years) who received either MF59Flu or HDFlu. Of these, 32 participants were excluded from the current study due to not receiving the vaccination (n = 9), incomplete study visits for blood draws (n = 7), or insufficient PBMCs for the assays (n = 16). The remaining cohort (n = 218) consisted of 106 MF59Flu recipients and 112 HDFlu recipients (**Table 1**, **Figure 1**). Both vaccine groups (MF59Flu and HDFlu) were comparable in demographic characteristics, including sex, age, ethnicity, race, and BMI (**Table 1**).

**Figure 1.**
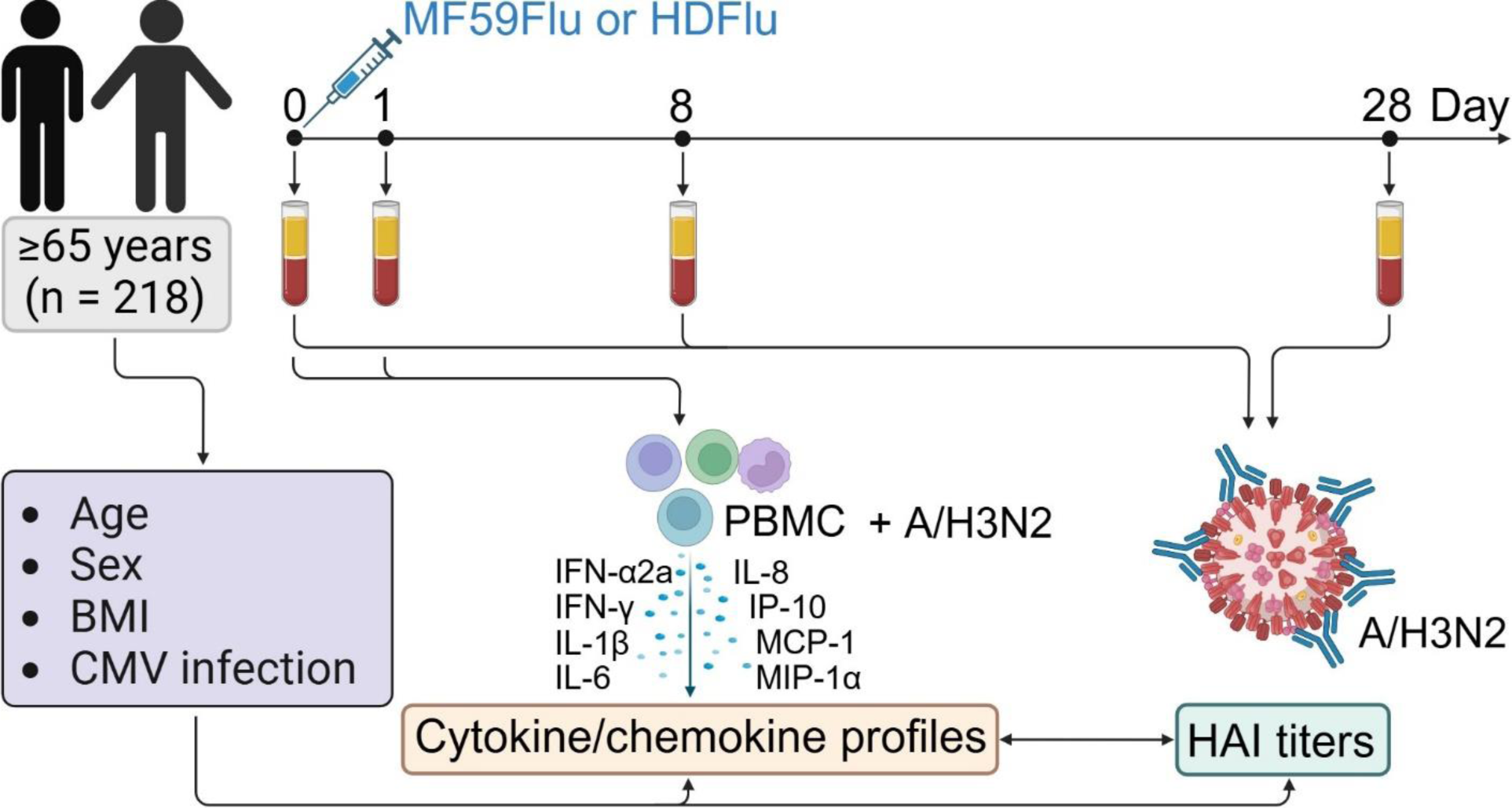
Study design. The study cohort included 106 recipients of the MF59-adjuvanted influenza vaccine (MF59Flu) and 112 recipients of the high-dose influenza vaccine (HDFlu). Blood samples were collected prior to vaccination (Day 0) and on Days 1, 8, and 28 post-vaccination. Peripheral blood mononuclear cells (PBMCs) were isolated from the Day 0 and Day 1 blood samples and *in vitro* stimulated with live influenza A/H3N2 virus. Production of eight pro-inflammatory cytokines and chemokines (IFN-α2a, IFN-γ, IL-1β, IL-6, IL-8, IP-10, MCP-1, MIP-1α) were quantified in the cell culture supernatants using an MSD multiplex assay. Additionally, serum antibody titers against the A/H3N2 virus were measured in sera collected at Days 0, 8, and 28 using the hemagglutination inhibition (HAI) assay. Associations between cytokine/chemokine responses and HAI titers were examined. The impact of key demographic and clinical factors, including age, sex, body mass index (BMI), and cytomegalovirus (CMV) infection status on the cytokine and chemokine profiles was assessed. Created in BioRender.com.

**Table 1.** Demographic characteristics of study cohort.

| Characteristics | MF59Flu (n = 106) | HDFlu (n = 112) | Total (n = 218) | *p* value |
| --- | --- | --- | --- | --- |
| **Sex** |  |  |  | 0.428<sup>a</sup> |
| Male (%) | 38 (36.8%) | 46 (41.1%) | 84 (38.5%) |  |
| Female (%) | 68 (64.2%) | 66 (58.9%) | 134 (61.5%) |  |
| **Age (years)** |  |  |  | 0.462<sup>b</sup> |
| Median (Q1, Q3) | 72 (68.1, 76.6) | 71.1 (67.7, 75.4) | 71.6 (67.9, 76) |  |
| **Ethnicity** |  |  |  | 0.956<sup>a</sup> |
| Non-Hispanic nor Latino | 104 (98.1%) | 110 (98.2%) | 214 (98.2%) |  |
| Hispanic or Latino | 2 (1.9%) | 2 (1.8%) | 4 (1.8%) |  |
| **Race** |  |  |  | 0.303<sup>a</sup> |
| Black or African American | 1 (0.9 %) | 0 | 1 (0.5%) |  |
| White | 105 (99.1%) | 112 (100%) | 217 (99.5%) |  |
| **BMI** |  |  |  | 0.429<sup>b</sup> |
| Median (Q1, Q3) | 28.45 (25.26, 31.57) | 27.99 (25.05, 31.18) | 28.28 (25.13, 31.51) |  |
| **CMV infection** |  |  |  |  |
| Negative |  |  | 91 |  |
| Equivocal |  |  | 2 |  |
| Positive |  |  | 125 |  |

<sup>a</sup>Pearson's Chi-squared test, <sup>b</sup>Wilcoxon rank sum test.

### 3.2. PBMCs from MF59Flu and HDFlu groups generate comparable levels of cytokines and chemokines

PBMCs sampled at Day 0 and Day 1 were incubated with live A/H3N2 virus overnight and the concentrations of eight cytokines and chemokines (IFN-α2a, IFN-γ, IP-10, MCP-1, MIP-1α, IL-1β, IL-6, and IL-8) were measured in the cell culture supernatant. Under unstimulated conditions, PBMCs from both Day 0 and Day 1 secreted these analytes at background levels (**Supplementary Figure S1**). However, upon stimulation with A/H3N2 virus, the concentration of these analytes increased over 10-fold (**Supplementary Figure S1**). Despite this robust response to viral stimulation, analyte concentrations were comparable between MF59Flu and HDFlu groups at either Day 0 or Day 1 (**Supplementary Figure S2**). We also did not observe any influence of sex on the change (Day 1 – Day 0) of analyte concentrations after vaccination **(Supplementary Figure S3)**.

### 3.3. IL-8 concentration is associated with age

At Day 0, there were no significant correlations between age and the concentrations of analytes (*p* >0.05) (**Figure 2**). However, age was positively correlated with IL-8 concentration at Day 1 (r = 0.24, *p* = 0.00028), as well as the change (Day 1 – Day 0) in IL-8 concentration (r = 0.16, *p* = 0.021) (**Figure 2A**). Additionally, age was positively correlated with IL-1β concentrations at Day 1 (r = 0.14, *p* = 0.042), but not the change in IL-1β concentration after vaccination (r = 0.097, *p* = 0.15) (**Figure 2B**). No other significant correlations were found between age and the concentrations of the remaining analytes (**Figure 2**).

**Figure 2.**
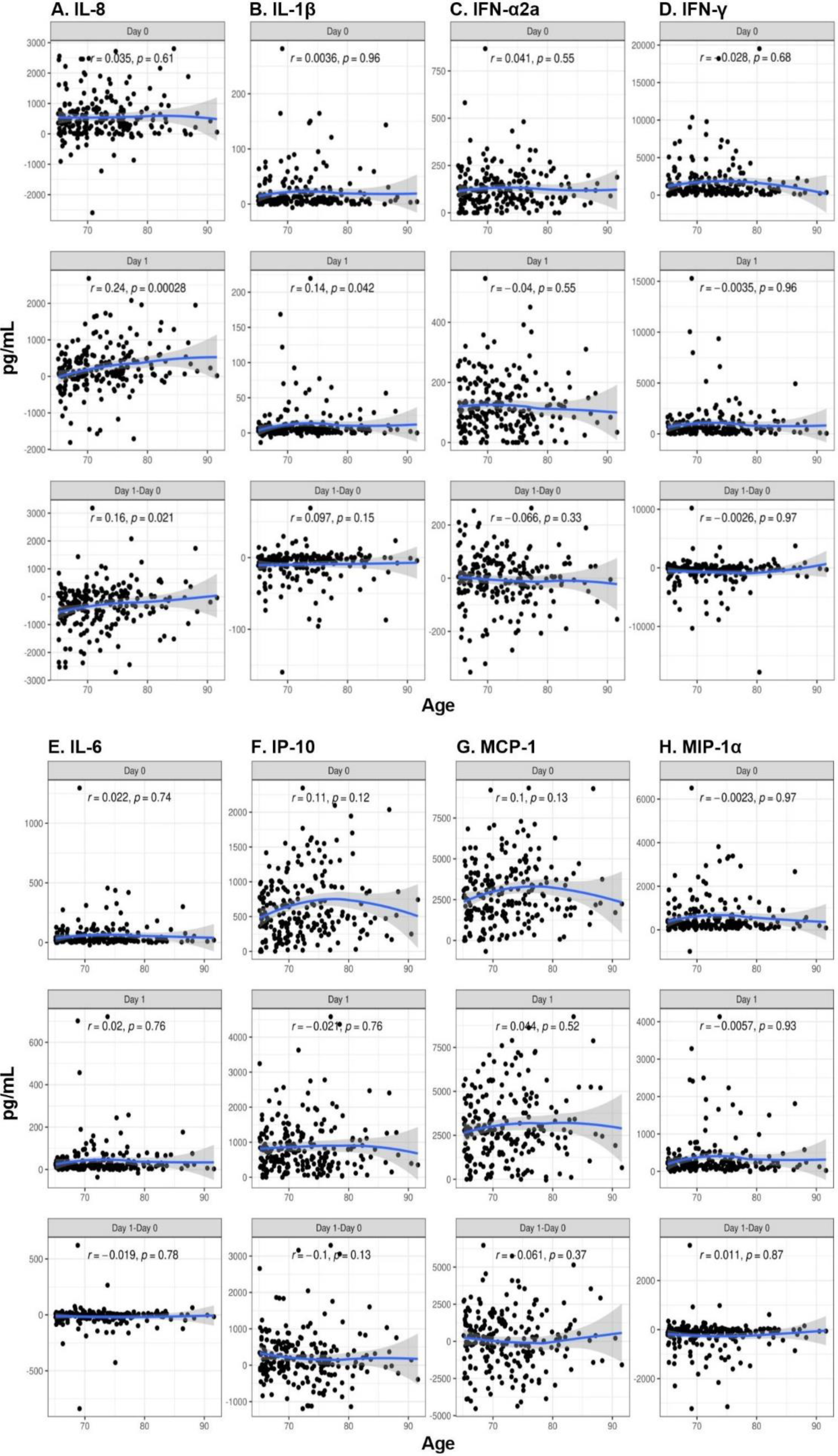
Positive correlation between age and IL-8. At Day 0, there was no significant correlation between age and IL-8 concentration (r = 0.035, *p* = 0.61, Figure 2A). However, age was positively correlated with IL-8 level at Day 1 (r = 0.24, *p* = 0.00028) and the change in IL-8 level from Day 1 to Day 0 (r = 0.16, *p* = 0.021). The correlations between age and the change in the concentration of other analytes were not significant (B-H).

### 3.4. Associations between HAI and cytokine or chemokine secretion

We measured HAI titers in serum samples collected at Days 0, 8, and 28, and examined the correlation between these titers and the change (Day 1 – Day 0) in analyte concentrations (**Figure 3, Supplementary Figure S4**). IP-10 exhibited a positive correlation with HAI titer (r = 0.16, p = 0.024) at Day 0, but not at Day 1. The change in IL-8 concentration did not significantly correlate with the HAI titers at Day 0 (r = −0.037, *p* = 0.59) although a negative trend was noticed at Day 8 (r = −0.13, *p* = 0.056). A significant negative association was observed between the decrease in IL-8 concentration at Day 1 compared to Day 0 and HAI titer at Day 28 (r = −0.15, *p* = 0.0026) (**Figure 3**). No significant correlations were found between changes in the concentration of other analytes and HAI titer (**Supplementary Figure S4**).

**Figure 3.**
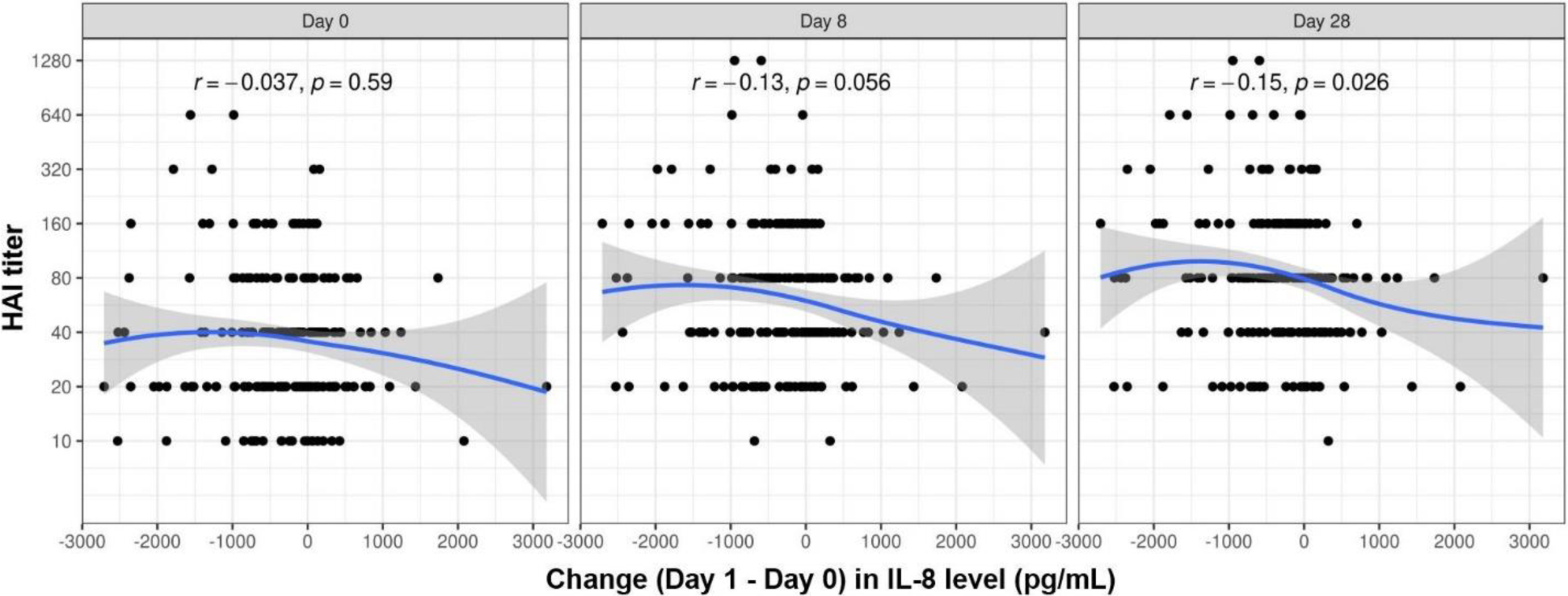
Negative correlation between the change in IL-8 secretion level and the HAI titers at Day 0, Day 8, and Day 28. Although there was a trend of negative correlation between the changes in IL-8 level from Day 0 to Day 1 and HAI titer at Day 0 (r = −0.037) and Day 8 (r = −0.13), these correlations were not statistically significant (p >0.05). The negative correlation between the change in IL-8 and HAI titers at Day 28 were significant.

### 3.5. CMV infection and BMI did not influence cytokine/chemokine responses

Prior cytomegalovirus (CMV) infection, a recognized marker of immunosenescence, has been linked to diminished immune responses to influenza vaccination [31]. In our cohort, we measured CMV-specific IgG in serum to determine CMV serostatus, identifying 91 CMV seronegative and 125 CMV seropositive recipients, as well as 2 recipients with CMV equivocal results (neither negative nor positive). Regardless of vaccine type (MF59Flu or HDFlu), the concentrations of the cytokine and chemokine analytes were comparable between CMV seronegative and CMV seropositive recipients (**Supplementary Figure S5**).

Additionally, no significant correlation was observed between body mass index (BMI) and the concentration of the analytes on both Day 0 and Day 1 (**Supplementary Figure S6**).

## 4. Discussion

Impaired immune responses in older adults (≥65 years) have led to the recommendation to use of either MF59Flu or HDFlu for this population. While humoral (antibody) responses to these vaccines are well-documented, less is known about the cellular responses, particularly regarding cytokine and chemokine production. In this study, we aimed to characterize early cellular immune responses by analyzing cytokine and chemokine production in PBMCs isolated from recipients of MF59Flu and HDFlu before (Day 0) and one day after vaccination (Day 1). This approach provides insights into early immune activation and potential differences between these two vaccine formulations in older adults. Additionally, we examined how sex, age, BMI, and CMV infection status might influence cytokine and chemokine responses. Understanding how these factors affect immune responses could inform strategies to optimize vaccination efficacy in this vulnerable population.

MF59Flu and HDFlu enhance immune responses through distinct mechanisms. Specifically, MF59Flu utilizes an oil-in-water emulsion adjuvant MF59, which rapidly induces the generation of inflammatory cytokines and chemokines, recruiting multiple innate immune cells to the site of injection [32]. In contrast, HDFlu contains a four-fold increase in antigen, compared to standard-dose influenza vaccines, but does not include any adjuvants [33]. Despite these differences, our study revealed surprisingly similar cytokine and chemokine response profiles between the two vaccine groups (**Supplementary Figures S1-S3**). Moreover, these response profiles were not significantly influenced by key demographic and clinical factors, such as sex, BMI, and CMV infection status (**Supplementary Figures S3, S5, S6**). While this finding contradicted our initial hypothesis, it is consistent with previous findings of similar vaccine effectiveness between MF59Flu and HDFlu [3–7]. These results suggest a potential correlation between the early production of pro-inflammatory cytokines and chemokines and subsequent antibody responses. Future studies comparing the efficacy of novel vaccine candidates against current influenza vaccines could leverage cytokine and chemokine profiles as early biomarkers, measurable as soon as one day post-vaccination, to predict vaccine effectiveness.

Previous studies have attributed the impaired responses to influenza vaccination in older adults primarily to the functional declines of both B and T cells [34–36]. However, effective activation of these adaptive immune cells after vaccination requires a supportive cytokine and chemokine milieu, largely produced by innate immune cells. In this study, we observed a positive association between age and IL-8 secretion post-vaccination (**Figure 2**). More notably, the early change in IL-8 concentration from Day 0 to Day 1 was negatively associated with HAI titers at Day 28 (**Figure 3**), suggesting a potential link between aging and diminished antibody responses in older adults to influenza vaccination. Previous studies have also reported a decrease in serum levels of IL-8 following influenza vaccination or infection [23, 37, 38]. In line with these findings, we observed a decline in IL-8 levels secreted by PBMCs one day after vaccination (**Supporting Figures S1 and S2**). This consistent reduction in IL-8 suggests that suppressing IL-8 production may play a pivotal role in enhancing the efficacy influenza vaccines in older adults.

The strength of our study lies in its well-controlled design. Our study cohort, consisting of recipients of both MF59Flu and HDFlu, had comparable demographic characteristics, which helped minimize potential impacts of these factors on cytokine and chemokine responses. Indeed, we did not observe any significant impacts of demographic factors, such as sex or BMI, on cytokine and chemokine responses (**Supplementary Figures S5 and S6**). However, we also acknowledge several limitations. First, we focused on antibody responses against the influenza A/H3N2 virus due to its particularly detrimental effects on older adults, but antibody responses to other vaccine strains may differ. Therefore, future studies should extend these findings by evaluating humoral responses to all influenza strains included in the vaccine. Second, the source of IL-8 remains to be fully elucidated. While IL-8 can be produced by multiple immune cell types, our previous findings suggested that monocytes, in collaboration of activated platelets, may be the primary source of IL-8 [39]. Further studies are needed to identify the specific sources and cell subsets responsible for maintaining a high baseline IL-8 level in older adults and the subsequent reduction in IL-8 post-vaccination. Understanding these cellular mechanisms could provide new targets to enhance vaccine-induced immune responses to influenza in older adults.

In summary, this study characterized cytokine and chemokine responses in *in vitro* stimulated PBMCs from recipients of the MF59Flu and HDFlu vaccines. Our results revealed a positive association between age and IL-8 concentration, which was negatively correlated with antibody responses. Although the precise cellular source of IL-8 remains to be determined, our findings suggest a potential link between age and impaired immune responses in older adults to influenza vaccine. This connection could serve as a potential target for molecular interventions aimed at enhancing influenza vaccine effectiveness in this high-risk population.

## Author Contributions

HQQ analyzed data and wrote the first draft of the manuscript. IGO, GAP, and RBK were involved in the conceptualization of the study. KMG and DEG performed statistical analyses. RBK and GAP were involved in the funding acquisition and project administration. All authors edited, revised, and approved submitted version of the manuscript.

## Funding

This research was funded by the National Institute of Allergy and Infectious Diseases of the National Institutes of Health with grant number R01AI132348, the Center for Influenza Vaccine Research for High-Risk Populations (CIVR-HRP) with contract number 75N93019C00052 (CIVIC). The content of this study is solely the responsibility of the authors and does not necessarily represent the official views of the National Institute of Health.

## Data Availability Statement

All data included in this study are presented as figures and tables and available from the corresponding author upon reasonable request.

## Acknowledgments

We would like to thank all participants in this study. We are grateful to the financial support from NIAID and CIVR-HRP.

## Conflicts of Interest

Dr. Poland is the chair of a safety evaluation committee for novel investigational vaccine trials being conducted by Merck Research Laboratories. Dr. Poland provides consultative advice to AiZtech; Emergent Biosolutions; GlaxoSmithKline; Invivyd; Janssen Global Services, LLC; Merck & Co. Inc.; Moderna; Novavax; and Syneos Health. Dr. Poland holds patents related to vaccinia, influenza, and measles peptide vaccines. Dr. Poland is an adviser to the White House and World Health Organization on COVID-19 vaccines and monkeypox, respectively. Drs. Poland, Kennedy, and Ovsyannikova have received grant funding and royalties from ICW Ventures for preclinical studies on a peptide-based COVID-19 vaccine. Drs. Poland, Kennedy, and Ovsyannikova have a pending patent related to COVID-19 peptide-based vaccines. Dr. Kennedy also offers consultative advice on vaccine development to Merck & Co. and Sanofi Pasteur. These activities have been reviewed by the Mayo Clinic Conflict of Interest Review Board and are conducted in compliance with Mayo Clinic Conflict of Interest policies. Other co-authors declare no competing interests.

---

**Supplementary Figure S1.**
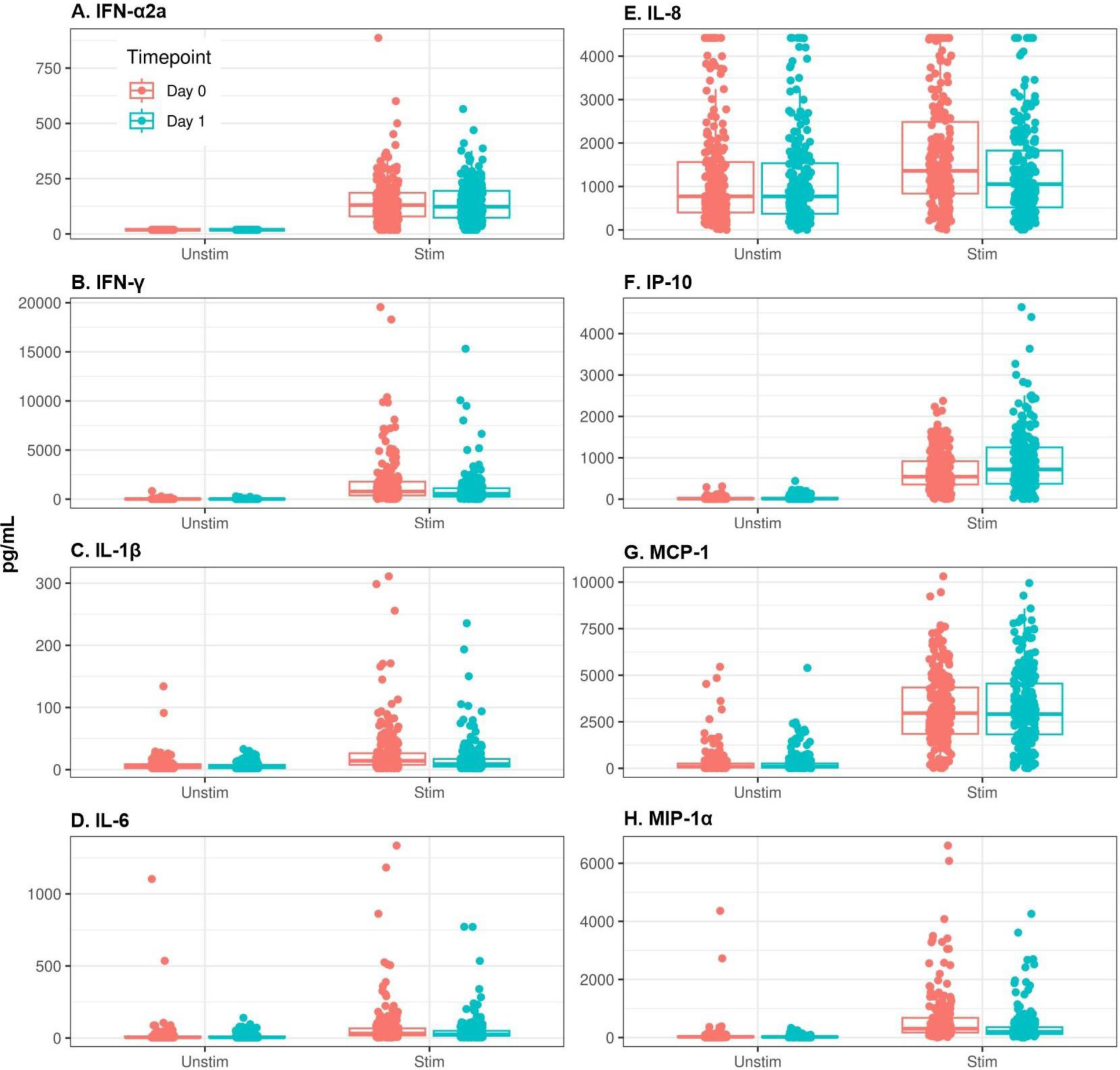
Significant changes in the production of cytokines and chemokines from *in vitro* stimulated PBMCs. Abbreviation: unstim = unstimulation (cell cultured in media); stim = H3N2 stimulation.

**Supplementary Figure S2.**
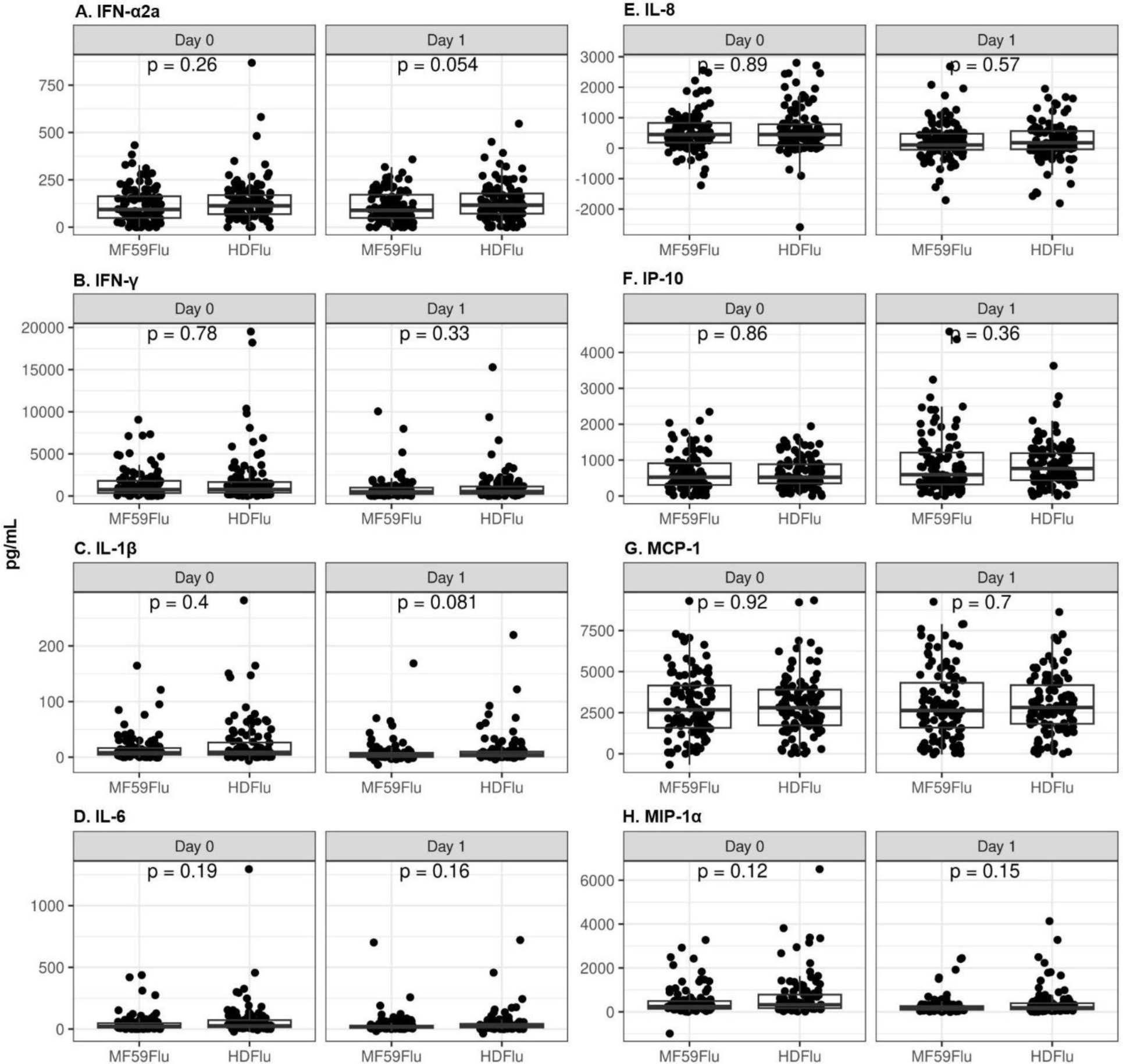
Nonsignificant influences of vaccine type (MF59Flu and HDFlu) in the production of cytokines and chemokines from in vitro stimulated PBMCs.

**Supplementary Figure S3.**
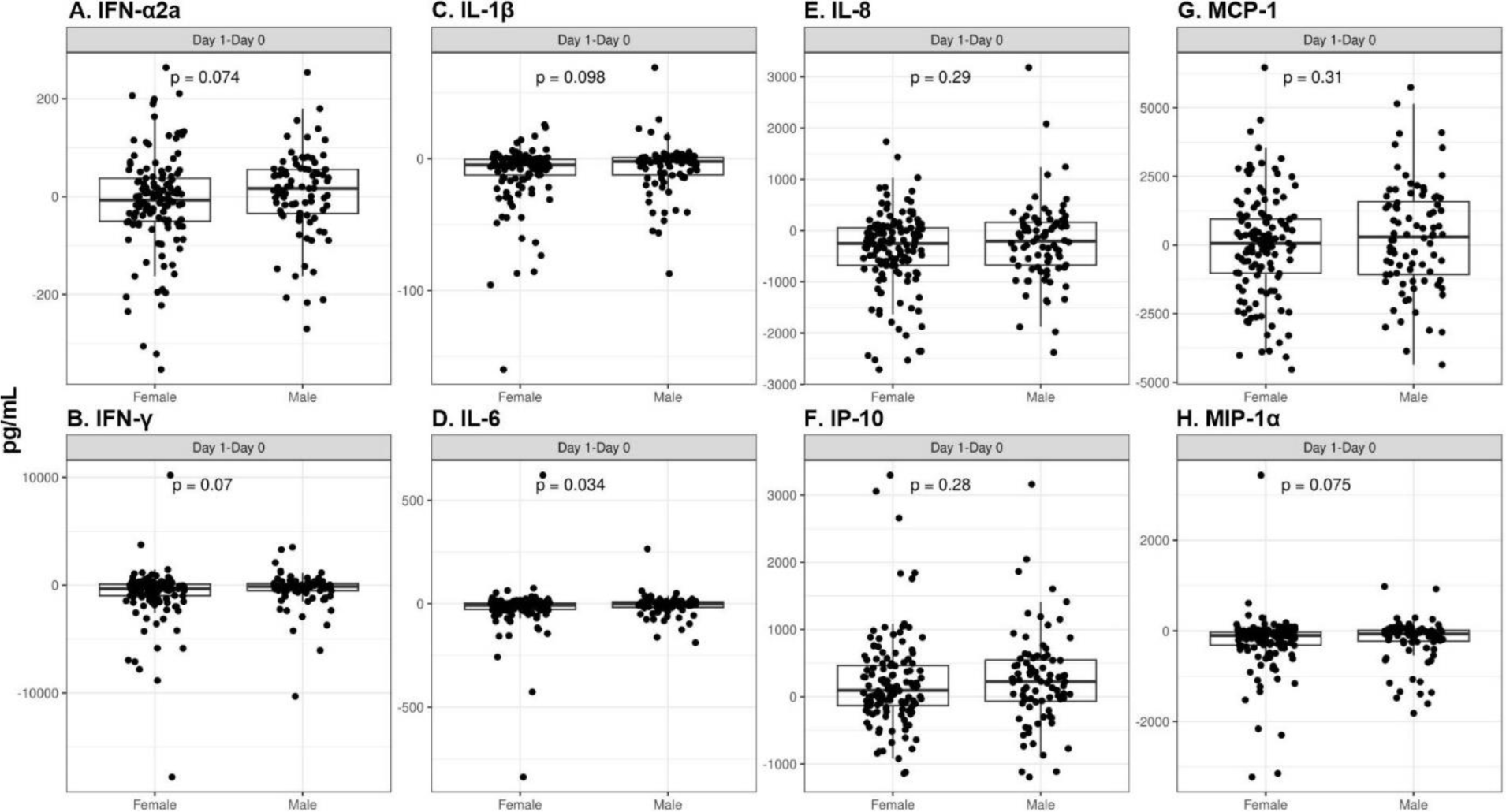
Nonsignificant influences of sex in the production of cytokines and chemokines from in vitro stimulated PBMCs.

**Supplementary Figure S4.**
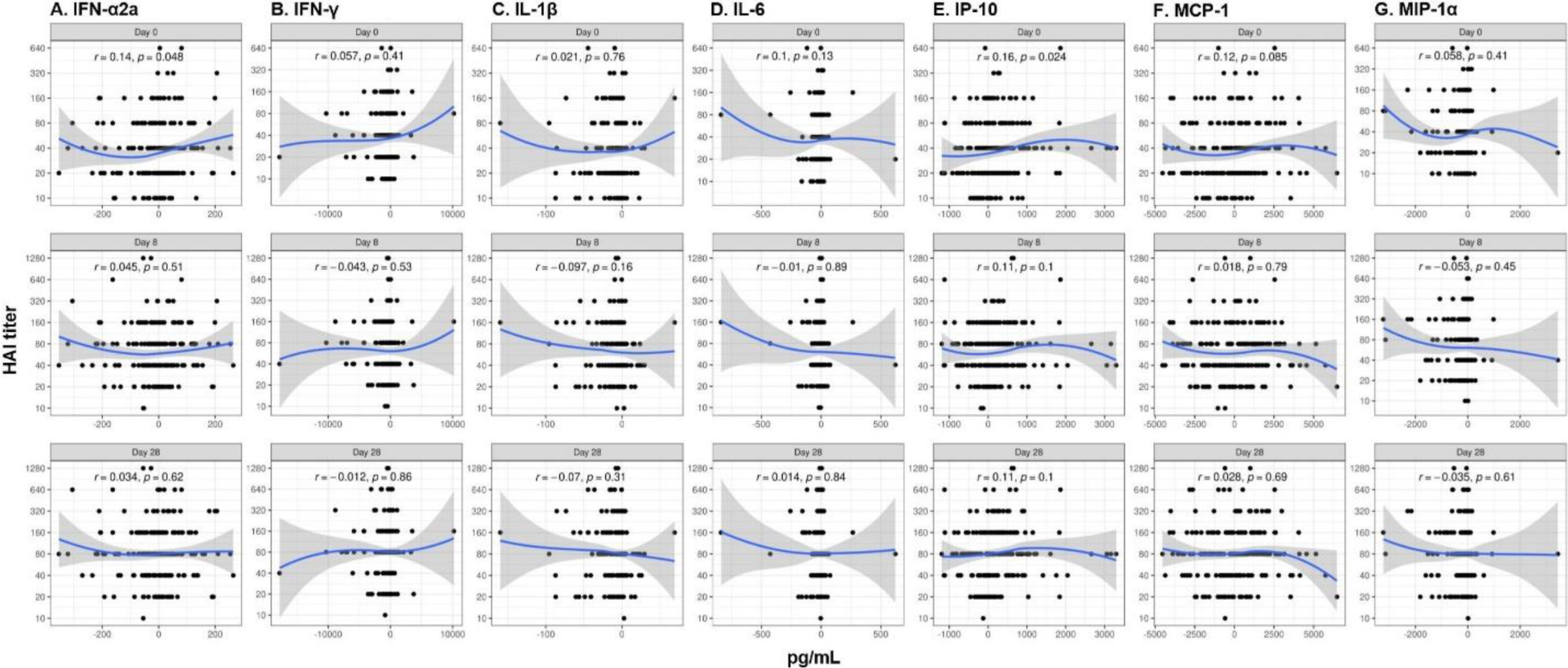
Nonsignificant correlations between the change in other cytokines and chemokines and the HAI titers at Day 0, Day 8, and Day 28.

**Supplementary Figure S5.**
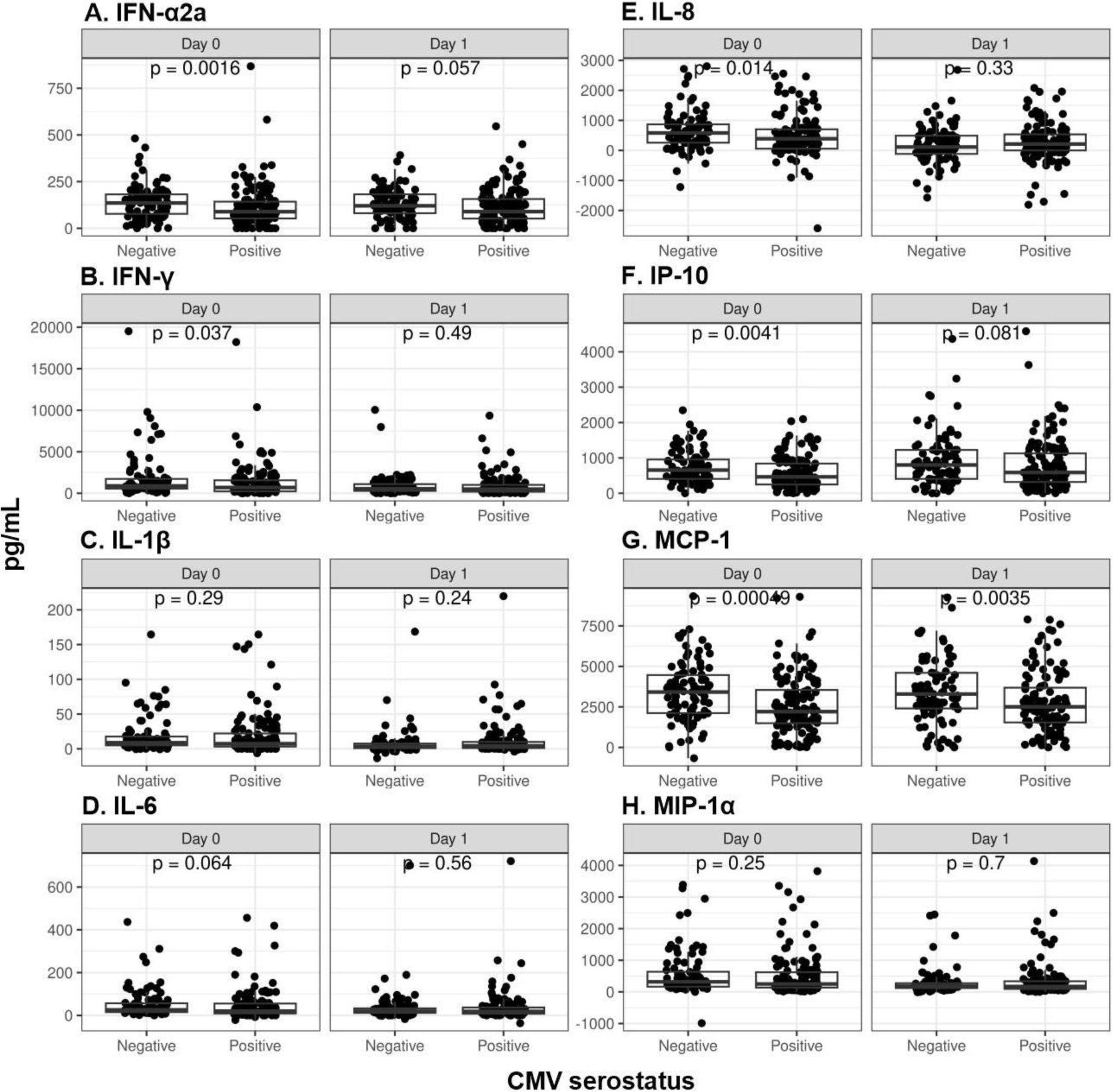
Nonsignificant impacts of CMV infection on the cytokine and chemokine responses.

**Supplementary Figure S6.**
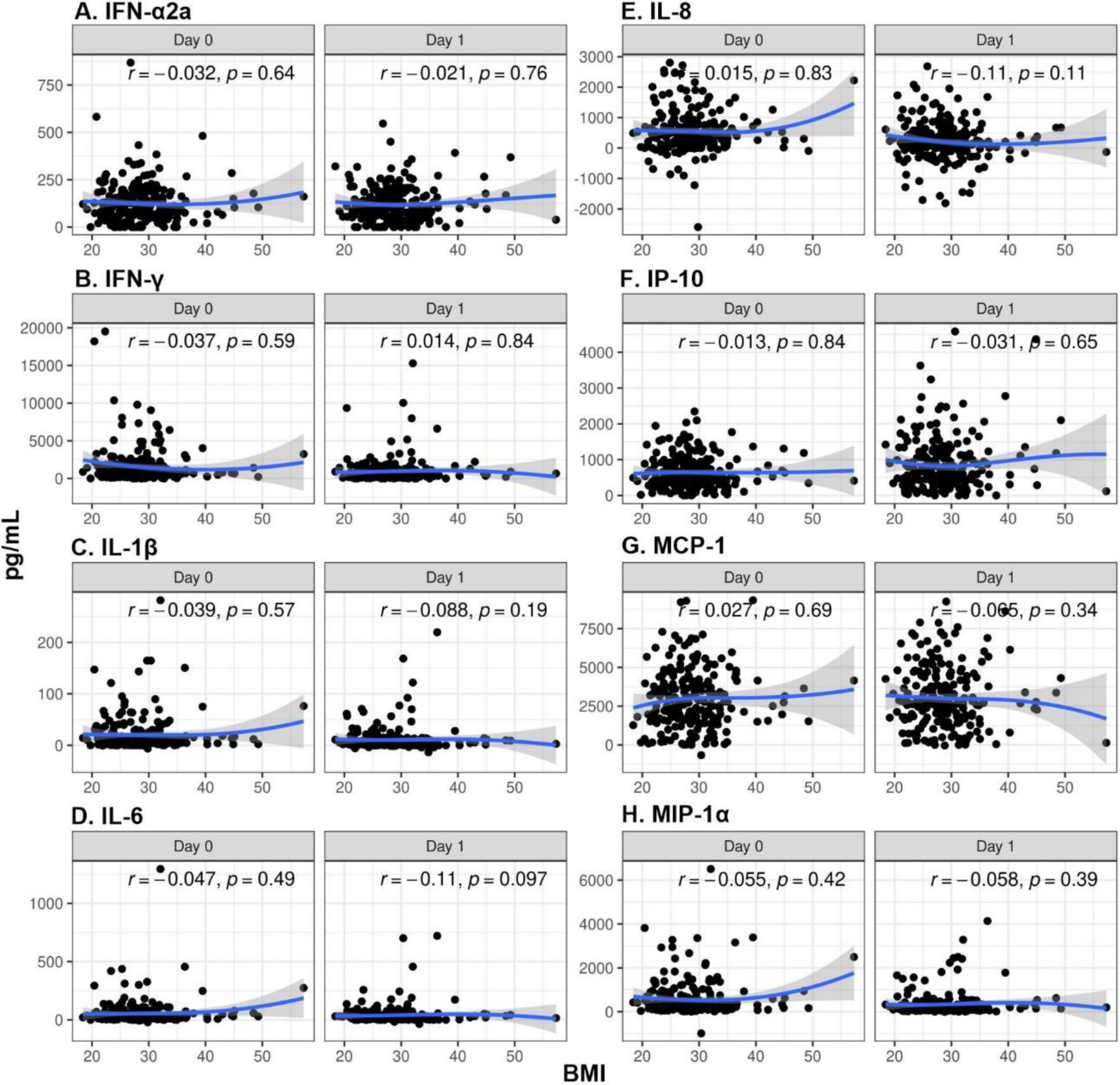
Nonsignificant correlations between BMI and the cytokine and chemokine responses.

